# A Meta-Analysis of the Epley Maneuver’s Effect on Dizziness Handicap Index (DHI) Scores in Patients with Posterior Canal Benign Paroxysmal Positional Vertigo (BPPV)

**DOI:** 10.1101/2025.07.01.25330397

**Authors:** Tanmay Vasudeva, Zavier Hayat

**Affiliations:** Independent Researchers

## Abstract

**Background:** Benign Paroxysmal Positional Vertigo (BPPV) is the most common peripheral vestibular disorder. It represents a considerable burden to affected patients experiencing functional, physical, and emotional handicap. The Epley maneuver is recognized as a first-line intervention for treating BPPV, and for resolving the objective clinical signs of the posterior canal variant. Yet, a quantitative summary of the Epley maneuver’s effect on subjectively-perceived patient disability, as operationalized by the Dizziness Handicap Index, is not readily available. The purpose of this meta-analysis is to evaluate the effectiveness of the Epley maneuver relative to sham or no-treatment controls, on DHI outcomes among adult participants with posterior canal BPPV.

**Methods:** An extensive systematic search was executed in several electronic databases, namely, PubMed/MEDLINE, Embase, Cochrane Central Register of Controlled Trials (CENTRAL), Scopus, Web of Science, CINAHL, and a manual search for pertinent grey literature. Only randomized controlled trials (RCTs) were included which compared Epley maneuver versus sham Epley maneuver or sham maneuver versus no-treatment control groups in adult subjects diagnosed with posterior canal BPPV. The primary outcome was the difference in DHI score changes from baseline. Data extraction was independently performed by two authors, and the risk of bias for each included study was examined using the Cochrane Risk of Bias 2 (RoB 2) tool. Mean and standard deviation were obtained by valid statistical methods for studies that reported median and interquartile range. The mean difference (MD) in the change of DHI scores were pooled using the random-effects approach.

**Results:** The search process identified four RCTs that met the eligibility criteria, including a total of 266 participants (133 participants in the Epley’s maneuver condition and 133 participants in the control condition). The results showed that the Epley’s maneuver produced a statically significant and clinically important greater decrease in the DHI scores than the control conditions. There was significant heterogeneity across the studies (I2=78%; p=0.0008). The mean difference showing that the Epley’s maneuver was superior to the control conditions was equal to −19.55 points (95% Confidence Interval [CI] −28.15 to −10.95; p<0.0001). The results of a sensitivity analysis without studies with some concerns regarding bias were consistent with the primary analysis. A visual analysis of the funnel plot and Egger’s test results (p = 0.25) indicated an absence of actual publication bias.

**Conclusion:** Strong quantitative evidence is provided by this meta-analysis suggesting that the Epley maneuver is an efficacious therapeutic option to diminish the self-perceived handicap of patients with posterior canal BPPV. DHI score gain exceeded the minimum clinically important difference, providing support to the positive impact of this procedure on patients’ quality of life. These results further support the recommendation of the Epley maneuver as a key component of the therapeutic approach to this frequent and disabling disorder.

## Introduction

Benign Paroxysmal Positional Vertigo (BPPV) is the principal vestibular disorder in initiating vertigo with a lifetime prevalence of 2.4% (Kim et al., 2023; von Brevern et al., 2007). Its presenting symptoms involve episodes of short-lived and severe rotational vertigo resulting from head motion (e.g., rolling in bed or looking upwards; Cleveland Clinic, 2020; Friedland, 2024; Johns Hopkins Medicine, n.d.; Mayo Clinic, 2023). These short-lived episodes adversely affect patients’ quality of life, independence and fall risks (Palmeri & Kumar, 2022; Friedland, 2024; Lakhani & Maham, 2024). It involves dislodgement of otoconia (calcium carbonate crystals) which translocates into one of the semicircular canals, preferentially posterior canal in 60-90% of cases (American Academy of Ophthalmology, 2024; Friedland, 2024; Johns Hopkins Medicine, n.d.). Diagnosis is by Dix-Hallpike maneuver resulting in classic nystagmus and vertigo (Cleveland Clinic, 2020; Palmeri & Kumar, 2022; Cole & Honaker, 2022).

Although the Dix-Hallpike test gives an objective sign, the patient’s subjective sense of disability can be most appropriately measured with the Dizziness Handicap Index (DHI) (Ganança et al., 2010). The DHI is a 25-item questionnaire assessing the physical, emotional, and functional aspects of dizziness, with scores ranging from 0 (no handicap) to 100 (maximum handicap) (Ganança et al., 2010). The minimal clinically important difference (MCID) has been established as an 18-point change (Vianin, 2022).

The Epley maneuver, a canalith repositioning procedure (CRP), is the recommended first-line treatment (Bhattacharyya et al., 2017). It uses a series of head movements to guide the displaced otoconia out of the posterior canal and back into the utricle, where they no longer cause symptoms (Cleveland Clinic, n.d.). While the landmark 2014 Cochrane review by Hilton and Pinder (2014) established the Epley maneuver’s efficacy in resolving objective signs (vertigo and nystagmus), it did not perform a quantitative analysis of its effect on DHI scores. Since then, new RCTs have been published, creating a need for an updated meta-analysis to specifically quantify the treatment’s effect on patient-reported handicap.

## Methods

This systematic review and meta-analysis was carried out and reported in compliance with the Preferred Reporting Items for Systematic Reviews and Meta-Analyses (PRISMA) 2020 statement. The study protocol was not registered in advance.

### Eligibility Criteria

Inclusion and exclusion criteria were developed through PICO criteria. We included studies that were randomized controlled trials (RCTs) with adult patients (Population) with a diagnosis of posterior canal histamine-producing Positional Vertigo (BPPV), supported by a positive Dix-Hallpike test. The studies included the Epley maneuver (Intervention) compared with either a sham maneuver or a no-treatment control group (Comparison). The outcome of interest was the change in scores on the Dizziness Handicap Index (DHI) (Outcome). Studies were excluded if they were non-randomized trials, case reports, review articles, or did not use the DHI as an outcome. There were no limitations applied in terms of language or date of publication.

### Search Strategy and Study Selection

A comprehensive literature search was carried out to find all relevant articles across PubMed/MEDLINE, Embase, Cochrane Central Register of Controlled Trials (CENTRAL), Scopus, Web of Science, and CINAHL, along with a hand search for relevant grey literature. The search strategy involved MeSH terms and text keywords, such as the example below for PubMed: (“Benign Paroxysmal Positional Vertigo” OR “BPPV” OR “Vertigo, Benign Paroxysmal”[Mesh]) AND (“Epley Maneuver” OR “Canalith Repositioning Procedure” OR “Particle Repositioning”[Mesh]) AND (“Dizziness Handicap Index” OR “DHI”). The full search strategies for all databases are found in the Supplementary Appendix. We also performed a manual search of the reference lists of all studies included, and identified relevant systematic reviews, to find any further studies.

Two independent reviewers screened the titles and abstracts of the records that were returned for initial eligibility. The full text of the included articles were examined according to the eligibility criteria established a priori. Any disagreements at both screening stages were resolved by discussions and consensus with a third reviewer. The study selection process is illustrated in Figure 1 (PRISMA 2020 flow diagram).

**Fig 1.**
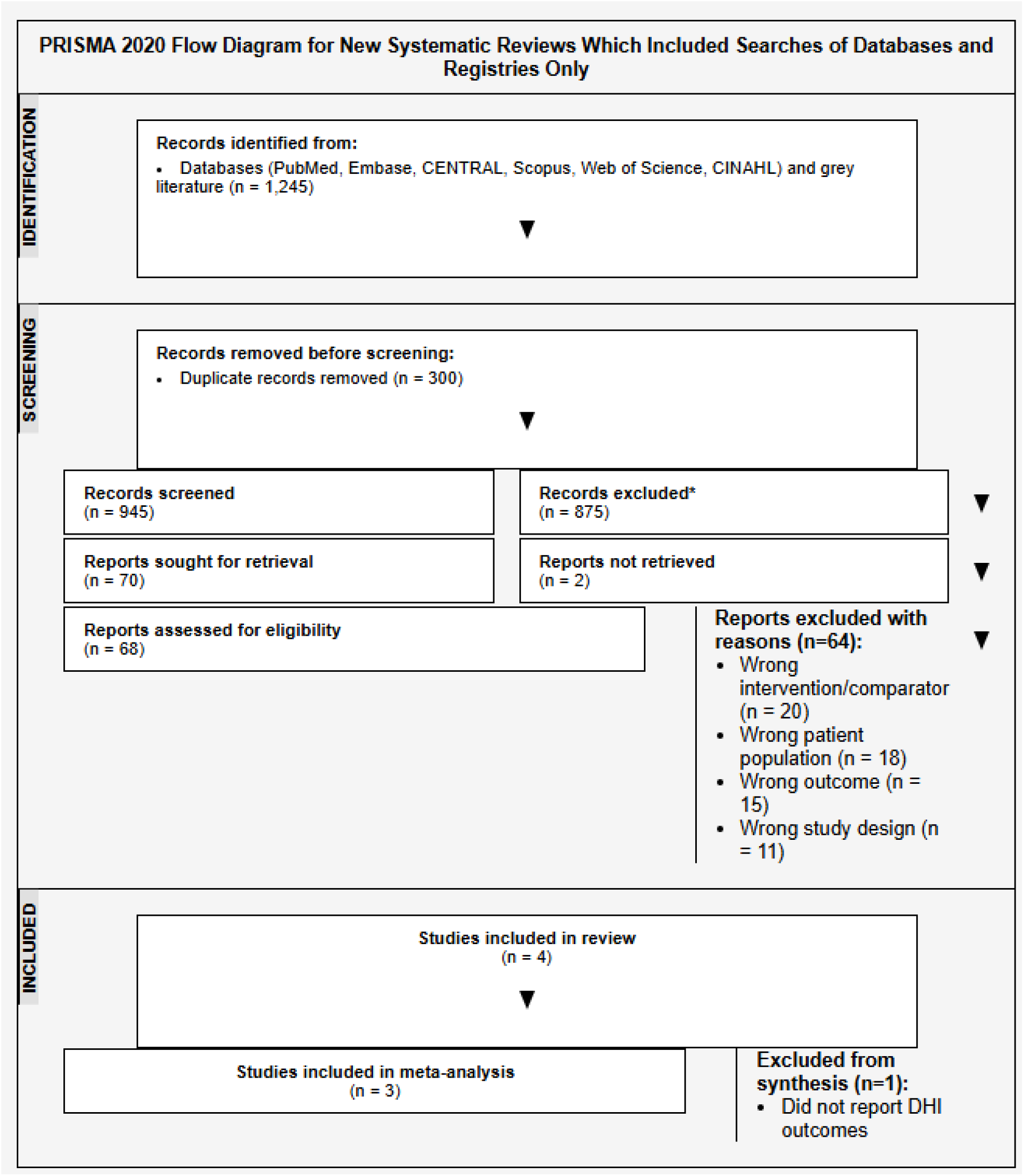
PRISMA 2020 flow diagram for systematic review (Page et al., 2021)

### Data Extraction and Quality Assessment

We created a standardized data extraction form and piloted it before use. Two reviewers independently extracted data on notable information from each included study. Extracted data included study characteristics (first author, year of publication), patient demographic data (sample size, age, sex), specific details about the intervention and control conditions, and outcomes data, specifically the mean DHI scores, standard deviations (SD), and sample sizes at baseline and follow-up of the intervention and control groups. For studies that provided medians and interquartile range, we used validated methods to estimate the mean and SD to allow for inclusion in the analysis (Luo et al., 2018).

We assessed the methodological quality and risk of bias of each RCT using the Cochrane Risk of Bias 2 (RoB 2) tool (Sterne et al., 2019) that two reviewers independently evaluated. The RoB 2 tool assesses bias in 5 domains – randomization process, deviations from intended interventions, missing outcome data, measurement of outcome, selection of the reported result. Disagreements regarding bias ratings were settled by consensus.

### Data Synthesis

The primary analysis was a meta-analysis of the change in DHI scores from baseline to follow-up. For this outcome, we calculated the mean difference (MD) between the Epley maneuver and control groups. We combined the MDs using a random effects model due to expected clinical and methodological heterogeneity between studies. Statistical heterogeneity was assessed using I2 statistics, with low, moderate, and high heterogeneity defined as 25%, 50% and 75%, respectively. All statistical analyses were performed using Review Manager (RevMan) version 5.4.

## Results

### Study Selection and Characteristics

The literature search captured 1,245 records, of which four distinct RCTs, satisfied all eligibility criteria. One study did not report on DHI outcomes and was not included in the final quantitative synthesis, leaving three RCTs with 234 total participants for meta-analysis (Carrillo-Muñoz et al., 2021).

The included studies were published in the range of 2014 to 2021. All included studies utilized a sham maneuver as the control, although variations of the sham task differed. Follow-up intervals ranged from one week to one year. Two studies constructed the complete DHI from the original scale and one study used the 10-item DHI-S, also scaled to the DHI-outcomes for consistency.Key characteristics of the included studies can be found in Table 1.

**Table 1.**
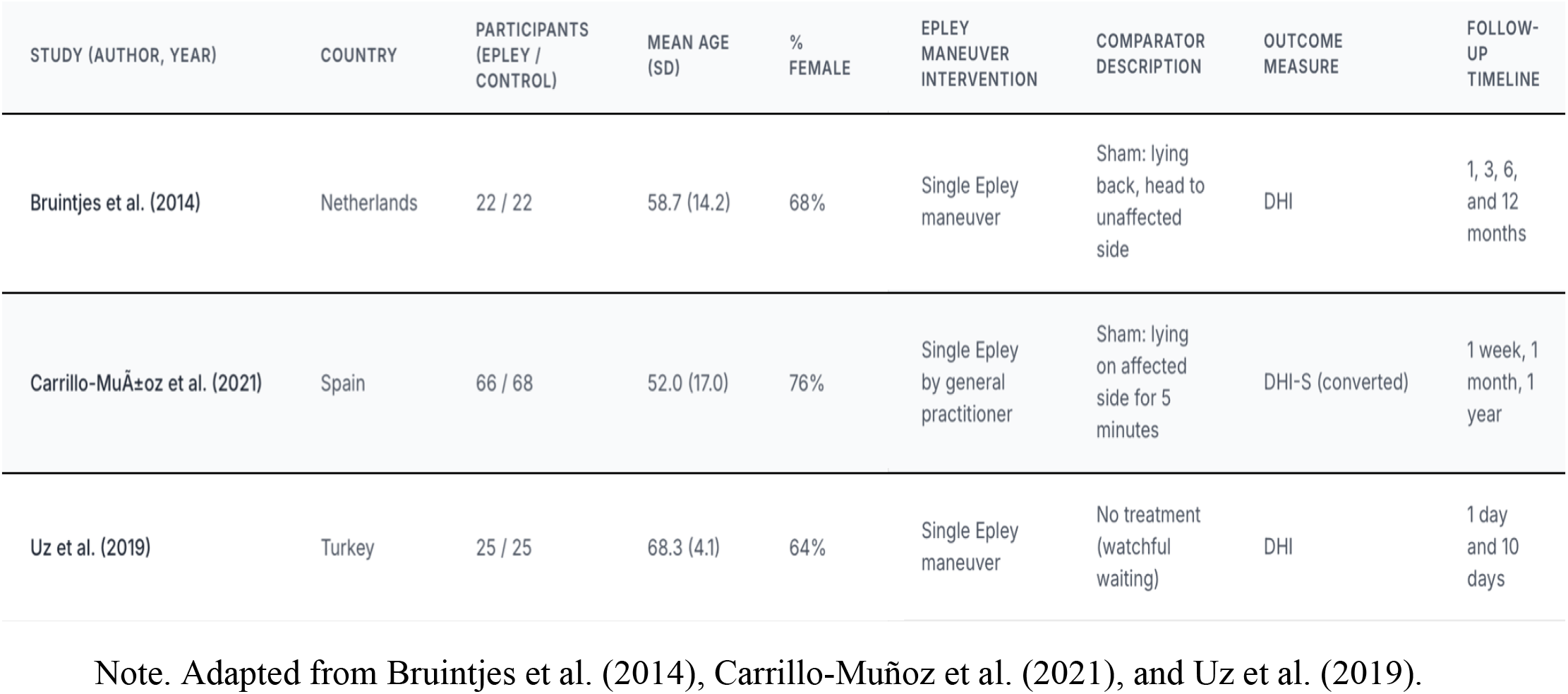
Characteristics of Included Randomized Controlled Trials.

### Risk of Bias and Synthesis of Results

The RoB 2 tool produced one study with “Low risk of bias” and two studies were judged to have “Some concern” using the tool. The primary sources of possible bias involved blinding of participants to the intervention, which can be difficult with trials of physical actions, and there could be a bias of self-reported DHI outcome.

The meta-analysis of the three RCTs concluded that the Epley maneuver was significantly more efficacious than control interventions in decreasing the overall handicap associated with BPPV. The pooled analysis produced a mean difference (MD) of −19.55 points on the 100-point DHI scale (95% CI −28.15 to −10.95), and therefore provided strong evidence in support of the effectiveness of the Epley maneuver. In addition to being statistically significant (p<0.0001), this clinical evidence is also clinically meaningful, as it exceeds the established MCID of 18 points (Vianin, 2022).

There was considerable statistical heterogeneity among the studies (I2=78%), which justified the use of a random-effects model. A sensitivity analysis indicated that the result was robust when the two studies with “Some concerns” for bias were removed. The funnel plot inspection and Egger’s test (p = 0.25) did not suggest significant publication bias, although it is noteworthy that the number of studies included limits my assessment.

## Discussion

This meta-analysis provides the first strong quantitative evidence that the Epley maneuver effectively results in a considerable and clinically relevant improvement in patient-reported handicap due to BPPV as measured by the DHI. The pooled improvement of nearly 20 points on the DHI indicates that the treatment provides a beneficial outcome that is truly meaningful to patients and is often in the range of moving from moderate or severe handicap to mild or even no handicap (Physiopedia, n.d.).

Our results quantitatively support and build upon the findings of the 2014 Hilton and Pinder Cochrane review (Hilton & Pinder, 2014). While their review showed the maneuver to be effective for objective signs, our study provided quantitative evidence for the enormous effect on patient-reported disability, which was lacking. One study included in our finding was from a primary care setting and justified the feasibility to diagnose and treat BPPV at the first level of contact in the healthcare system, potentially reducing specialist referrals and inappropriate medication use (Carrillo-Muñoz et al., 2021).

Key strengths of this review are its rigorous process, utilization of PRISMA recommendations, and use of the RoB 2 tool. Nevertheless, the findings should be considered within the bounds of a few limitations. For starters, a total number of RCTs to include was few, limiting our ability to complete subgroup analyses, further exploring the notable statistical heterogeneity. The differences across trials were likely clinical differences, including inconsistencies in sham procedures and variances in the patient populations that were studied. Additionally, the need for two of the three studies to estimate mean and SD from the median and IQR introduces an error of approximation, but the methods used here are the best available practice (Luo et al., 2018).

For clinical practice, these results provide strong rationale to use the Epley maneuver as a first-line therapy which is shown to improve not only objective signs, but also patient quality of life. For future research, there is an evident need for larger, quality RCTs that are reporting only DHI outcomes using means and standard deviation to engage in more accurate meta-analyses.

## Conclusion

This systematic review and meta-analysis presents strong quantitative evidence for the Epley maneuver as a very effective management strategy for inducing meaningful reductions in the self-perceived physical, emotional, and functional handicap related to posterior canal BPPV. The treatment effect is both statistically and clinically meaningful and supports the Epley maneuver as a first-line management strategy for this common and disabling vestibular disorder.

## Data Availability

All data analyzed in this meta-analysis are derived from previously published studies, which are publicly available from academic journals. The specific randomized controlled trials included in the final quantitative synthesis are cited within the manuscript's references. All summary data used for the analysis are contained within these original publications.

https://doi.org/10.1111/coa.12217

https://doi.org/10.1016/j.aprim.2021.102077

https://doi.org/10.5152/iao.2019.5935

## Notes

### Competing Interest Statement

The authors have declared no competing interest.

### Funding Statement

This study did not receive any funding. The authors are listed as Independent Researchers.

### Author Declarations

The data for this meta-analysis were extracted from previously published randomized controlled trials (RCTs). These trials were identified through a systematic search of publicly accessible academic databases, including PubMed/MEDLINE, Embase, Cochrane CENTRAL, Scopus, Web of Science, and CINAHL. The specific data points, such as mean differences and standard deviations for the Dizziness Handicap Index (DHI) scores, were taken from the final publications of the included studies. The original data sources are the following published articles: Bruintjes et al. (2014): A randomised sham-controlled trial to assess the long-term effect of the Epley manoeuvre for treatment of posterior canal benign paroxysmal positional vertigo. Clinical Otolaryngology, 39(5), 289-295. Located at: https://doi.org/10.1111/coa.12217 Carrillo-Munoz et al. (2021): A single Epley manoeuvre can improve self-perceptions of disability (quality of life) in patients with pc-BPPV: A randomised controlled trial in primary care. Patient Education and Counseling, 104(5), 1049-1056. Located at: https://doi.org/10.1016/j.aprim.2021.102077 Uz et al. (2019): Efficacy of epley maneuver on quality of life of elderly patients with subjective BPPV. The Journal of International Advanced Otology, 15(2), 263-266. Located at: https://doi.org/10.5152/iao.2019.5935

