## Supplemental Appendix 1 for "A Meta-Analysis of the Epley Maneuver’s Effect on Dizziness Handicap Index (DHI) Scores in Patients with Posterior Canal Benign Paroxysmal Positional Vertigo (BPPV)"

**Supplementary Appendix: Detailed Search Strategies**

This document provides the complete search strategies used for each electronic database, as referenced in the Methods section of the manuscript "A Meta-Analysis of the Epley Maneuver's Effect on Dizziness Handicap Index (DHI) Scores in Patients with Posterior Canal Benign Paroxysmal Positional Vertigo (BPPV)."

### 1. PubMed/MEDLINE

*Last searched: January 20, 2025*

| Search # | Query |
|---|---|
| **#1** | "Benign Paroxysmal Positional Vertigo"[Mesh] OR "Benign Paroxysmal Positional Vertigo"[tiab] OR "BPPV"[tiab] |
| **#2** | "Epley Maneuver"[Mesh] OR "Canalith Repositioning Procedure"[tiab] OR "Particle Repositioning Maneuver"[tiab] |
| **#3** | "Dizziness Handicap Index"[tiab] OR "DHI"[tiab] |
| **#4** | #1 AND #2 AND #3 |

### 2. Embase (via Ovid)

*Last searched: February 5, 2025*

| Search # | Query |
|---|---|
| **#1** | 'benign paroxysmal positional vertigo'/exp OR 'benign paroxysmal positional vertigo':ab,ti OR 'bppv':ab,ti |
| **#2** | 'epley maneuver'/exp OR 'canalith repositioning procedure':ab,ti OR 'particle repositioning maneuver':ab,ti |
| **#3** | 'dizziness handicap index':ab,ti OR 'dhi':ab,ti |
| **#4** | #1 AND #2 AND #3 |

### 3. Cochrane Central Register of Controlled Trials (CENTRAL)

*Last searched: February 28, 2025*

| Search # | Query |
|---|---|
| **#1** | [mh "Benign Paroxysmal Positional Vertigo"] OR "Benign Paroxysmal Positional Vertigo":ti,ab OR "BPPV":ti,ab |
| **#2** | [mh "Epley Maneuver"] OR "Canalith Repositioning Procedure":ti,ab OR "Particle Repositioning Maneuver":ti,ab |
| **#3** | "Dizziness Handicap Index":ti,ab OR "DHI":ti,ab |
| **#4** | #1 AND #2 AND #3 |

#### 4. Scopus

*Last searched: March 10, 2025*

( TITLE-ABS-KEY ( "Benign Paroxysmal Positional Vertigo" OR "BPPV" ) )
AND
( TITLE-ABS-KEY ( "Epley Maneuver" OR "Canalith Repositioning Procedure" OR "Particle Repositioning Maneuver" ) )
AND
( TITLE-ABS-KEY ( "Dizziness Handicap Index" OR "DHI" ) )
AND
( DOCTYPE ( ar ) )

#### 5. Web of Science (Core Collection)

*Last searched: April 2, 2025*

| Search # | Query |
|---|---|
| **#1** | TS=("Benign Paroxysmal Positional Vertigo" OR "BPPV") |
| **#2** | TS=("Epley Maneuver" OR "Canalith Repositioning Procedure" OR "Particle Repositioning Maneuver") |

| | |
|---|---|
| **#3** | TS=("Dizziness Handicap Index" OR "DHI") |
| **#4** | #1 AND #2 AND #3 |

#### 6. CINAHL (via EBSCO)

*Last searched: May 25, 2025*

| **Search #** | **Query** |
|---|---|
| **S1** | (MH "Benign Paroxysmal Positional Vertigo+") OR TI "Benign Paroxysmal Positional Vertigo" OR AB "Benign Paroxysmal Positional Vertigo" OR TI "BPPV" OR AB "BPPV" |
| **S2** | (MH "Epley Maneuver+") OR TI "Epley Maneuver" OR AB "Epley Maneuver" OR TI "Canalith Repositioning" OR AB "Canalith Repositioning" |
| **S3** | TI "Dizziness Handicap Index" OR AB "Dizziness Handicap Index" OR TI "DHI" OR AB "DHI" |
| **S4** | S1 AND S2 AND S3 |
